# Evaluation of Sample Pooling for Screening of SARS CoV-2

**DOI:** 10.1101/2020.06.10.20123398

**Authors:** Andargachew Mulu, Dawit Hailu Alemayehu, Fekadu Alemu, Dessalegn Abeje Tefera, Sinknesh Wolde, Gebeyehu Aseffa, Tamrayehu Seyoum, Meseret Habtamu, Alemseged Abdissa, Abebe Genetu Bayih, Getachew Tesfaye Beyene

**Affiliations:** Armauer Hansen Research Institute, P. O. Box 1005, Addis Ababa, Ethiopia

**Keywords:** RNA, nucleic acid, RT-PCR, Pooling, SARS CoV-2, COVID-19

## Abstract

**Background:** The coronavirus disease (COVID-19) pandemic has revealed the global public health importance of robust diagnostic testing. To overcome the challenge of nucleic acid (NA) extraction and testing kit availability efficient method is urgently needed.

**Objectives:** To establish an efficient, time and resource-saving and cost-effective methods, and to propose an *ad hoc* pooling approach for mass screening of SARS-CoV-2

**Methods:** Direct clinical sample and NA pooling approach was used for the standard reverse transcriptase polymerase chain reaction (RT-PCR) test of the SARS CoV-2 targeting the envelop (E) and open reading frame (ORF1ab) genomic region of the virus. In this approach, experimental pools were created using SARS CoV-2 positive clinical samples spiked with up to 9 negative samples prior to NA extraction step to have a final extraction volume of 200 μL (maximum dilution factor of 10). Viral NA was also subsequently extracted from each pool and tested using the SARS CoV-2 RT-PCR assay.

**Results:** We found that a single positive sample can be amplified and detected in pools of up to 7 samples depending on the ct value of the original sample, corresponding to high, medium, and low SARS CoV-2 viral copies/reaction. However, to minimize false negativity of the assay with pooling strategies and with unknown false negativity rate of the assay under validation, we recommend poling of 4 in 1 using the standard protocols of the assay, reagents and equipment. The predictive algorithm indicated a pooling ratio of 4 in 1 was expected to retain accuracy of the test irrespective of the ct value (relative RNA copy number) of the sample spiked and result in a 237% increase in testing efficiency.

**Conclusions:** The approaches showed its concept in easily customized and resource-saving manner and would allow expanding of current screening capacities and enable the expansion of detection in the community.

## Background

The coronavirus induced disease 2019 (COVID-19) pandemic has revealed the global public health importance of robust diagnostic testing to differentiate severe acute respiratory syndrome coronavirus 2 (SARS CoV-2) from other routine respiratory infections and to guide appropriate public health and individual clinical management [1]. Detecting carriers of the virus at various population levels is fundamental to response efforts. It ensures the quarantine of COVID-19 patients to prevent local community transmission, and more broadly informs national response team to take measures [2]. However, it remains uncertain whether there may have been community circulation of SARS CoV-2 prior to the identification of individuals with positive results through standard public health surveillance as detection and monitoring capacity is limited [3], and testing in Ethiopia is generally done on handful of facilities, while potentially infectious carriers at the community remain undiagnosed. Given the limited testing capacity available in Ethiopia, the decision to test is based on clinical and epidemiological factors and linked to an assessment of the likelihood of infection and testing of appropriate specimens from patients meeting the suspected case definition for COVID-19 is a priority for clinical management and outbreak control [3]. Thus, it is necessary to come up with new ways to efficiently and effectively use available resources.

Sample pooling (mixing of samples and testing at a single pool, and subsequent individual testing needed only if the pool tests positive) has been used as an attractive method for community monitoring of infectious diseases as it requires no additional training, equipment, or materials [4–6]. The key principles for successful application of group testing involve knowledge of the limit-of-detection, sensitivity, and specificity of the assay, and the prevalence of disease in a given population (7, 8). Here we have shown a proof-of-concept for direct clinical sample and NA pooling for the diagnosis of SARS CoV-2 in Ethiopia using the existing assay.

### Objective

To establish an efficient, time and resource-saving and cost-effective methods and to propose an *ad hoc* laboratory-based surveillance approach for mass screening of SARS-CoV-2

## Methods

### Design

The workflow comprises extraction of nucleic acid (NA) from individual respiratory samples (Nasopharyngeal and oropharyngeal swabs in viral transport medium), pooling of these respiratory samples and NA extraction, and pooling of extracted NA samples in batches of 10 and conducting SARS-CoV-2 specific real-time RT-PCR using the Novel Coronavirus 2019-nCov PCR Kit-fluorescent PCR method of Da An Gene Co., Ltd, China which is used currently for the diagnosis of SARS CoV-2 in the country. Nucleic acid was extracted from 200 μL respiratory specimen using the NA extraction and Purification Reagent, DAAN Gene Co., Ltd, as recommended by the manufacturer (Da An Gene Co., Ltd, of Sun Yat-Sen University, China). All laboratory procedures (Sample processing-NA extraction and purification, master mix (MM) preparation, mixing of NA and MMs, amplification/detection and analysis) were performed according to the manual provided by the manufacturer (Da An Gene Co., Ltd) on an identical qRT-PCR on BioRad CFX96 Deep Well Real-Time System, BioRad Laboratories, Inc, Singapore and program. Change in cycle threshold (ct) value (which is defined as the cycle threshold value of a reaction when the fluorescence of a PCR product can be detected above the background signal) of positive sample were analyzed. Here, a low and high Ct value corresponds to the presence of higher and lower amounts of viral RNA, respectively. The assay targets E and ORF 1ab region of SARS CoV-2. With this assay, a positive SARS CoV-2 result is determined when both targets reach a defined Ct value of less than 40, along with defined Ct value of less than 32 and 40 for positive control and internal control, respectively.

### Pooling

We conducted the pooling in two arms (direct clinical samples arm and nucleic acid arm) and each reaction was done in triplicate.

First, we pooled direct clinical samples of previously known positive samples with low, medium and high ct values corresponding to low, medium, and high viral copy number, respectively up to 10 samples in 1 prior to NA extraction step (maximum dilution factor of 10) to a final extraction volume of 200 μL when combined with an increasing number of previously confirmed negative samples as indicated in **Table 1**. In this study, a positive sample with ct value ≤ 34 considered as low ct value, between [> 34 and ≤37] medium, and between [>37 and ≤40] is high. Then, NA was extracted from final pooled samples of 200μL with a final elution volume of 50 μL. From the eluate template NA,5 µl of eluate was mixed with 20 µl of the RT-qPCR reagent master mix to have a final volume of 25 µl reaction mixture. Then, change in ct value of the positive control, positive samples with low, medium and high ct value and the cycle when all tested with no ct value were analyzed. Finally, we analyzed the impact of pool testing in batches of 10 samples per pool.

**Table 1:** Direct clinical sample Pools tested for SARS-CoV-2 RNA

| Pooling proportion (Dilution) | Positive Sample volume (µl) | Pool of up to 9 negative samples (µl) |
| --- | --- | --- |
| 1 (Original) | 200 | 0 |
| 1:1 (2 in 1) | 100 | 100 |
| 1:2 (3 in 1) | 67 | 133 |
| 1:3 (4 in 1) | 50 | 150 |
| 1:4 (5 in 1) | 40 | 160 |
| 1:5 (6 in 1) | 34 | 166 |
| 1:6 (7 in 1) | 29 | 171 |
| 1:7 (8 in 1) | 25 | 175 |
| 1:8 (9 in 1) | 23 | 177 |
| 1:9 (10 in 1) | 20 | 180 |

Second, we pooled individual NA preparation extracted from 200μL of direct clinical samples in pools of 10 (dilution factor of 10) in a final elute volume of 50μL proportionally to have a final NA volume of 5μL as indicated above in **Table 1**. For detecting a single positive sample within a pool of negative nucleic acid extracts, we evaluated the ability of the standard qRT-PCR test under the protocol recommended by manufacture of the kits. Then, change in ct value of samples with low, medium and high ct value was analyzed.

## Results

To assess the pool testing strategy, the most efficient pool size was calculated using a web based application of pooling (https://www.chrisbilder.com/shiny). As per the key principles of pooling, the following assumptions with numeric parameters are considered: an experimental prevalence rate of SARS CoV-2 in Ethiopia to be 0.05% (as observed positive rate within the tested individuals is reaching to 0.66% in the last 5 weeks), a two-stage pooling in a range of pool sizes 3–10 samples, an assay limit of detection (LOD) of 2.5 RNA copies/μL of reaction, an assay sensitivity of 98% – 100% and an assay specificity of 100%. With these calculations, a pool size of 4 samples predicted and would provide the largest reduction in the expected number of tests of 60% when compared to testing clinical samples separately (Table 2).

**Table 2:** Comparison of optimal sample pool size and rates of test efficiency* *Calculated using Shiny application of pooling strategy available at http://www.chrisbilder.com/shiny with the specified key principles of pooling indicated above

| Optimal sample pool size | Expected increase in testing Efficiency (5) | Reduction in Expected number of tests (%) |
| --- | --- | --- |
| 3 | 181 | 59 |
| 4 | 237 | 60 |
| 5 | 303 | 61 |
| 6 | 378 | 63 |
| 7 | 461 | 65 |
| 8 | 549 | 68 |
| 9 | 643 | 71 |
| 10 | 741 | 741 |

With our pooling strategy, we were able to detect SARS CoV-2 positives samples in pooling up to 8 in 1 which tested positive in individual RT-PCR (Figure 1 and 2). The results showed that pooled samples were positive within a range of 0 Ct to 6.75 Ct value difference from the original samples. Briefly, 54 pools of 6 specimens, each containing one positive sample were group tested. Of these, the SARS CoV-2 positive samples with original low ct value (high viral copy number) were within a range of Ct values from 28.61 to 32.97 for nucleocapsid (N) gene (Figure 1a) and from 30.77 to 37.53 for the open reading frame (ORF)1ab genes (Figure 1b) in 10 in 1 pool. Figure 1a and b shows the change in ct value of a high ct value positive sample spiked with up to 9 negative samples that is up to a dilution factor of 10 for the two targets, and ORF1ab genes.

**Figure 1:**
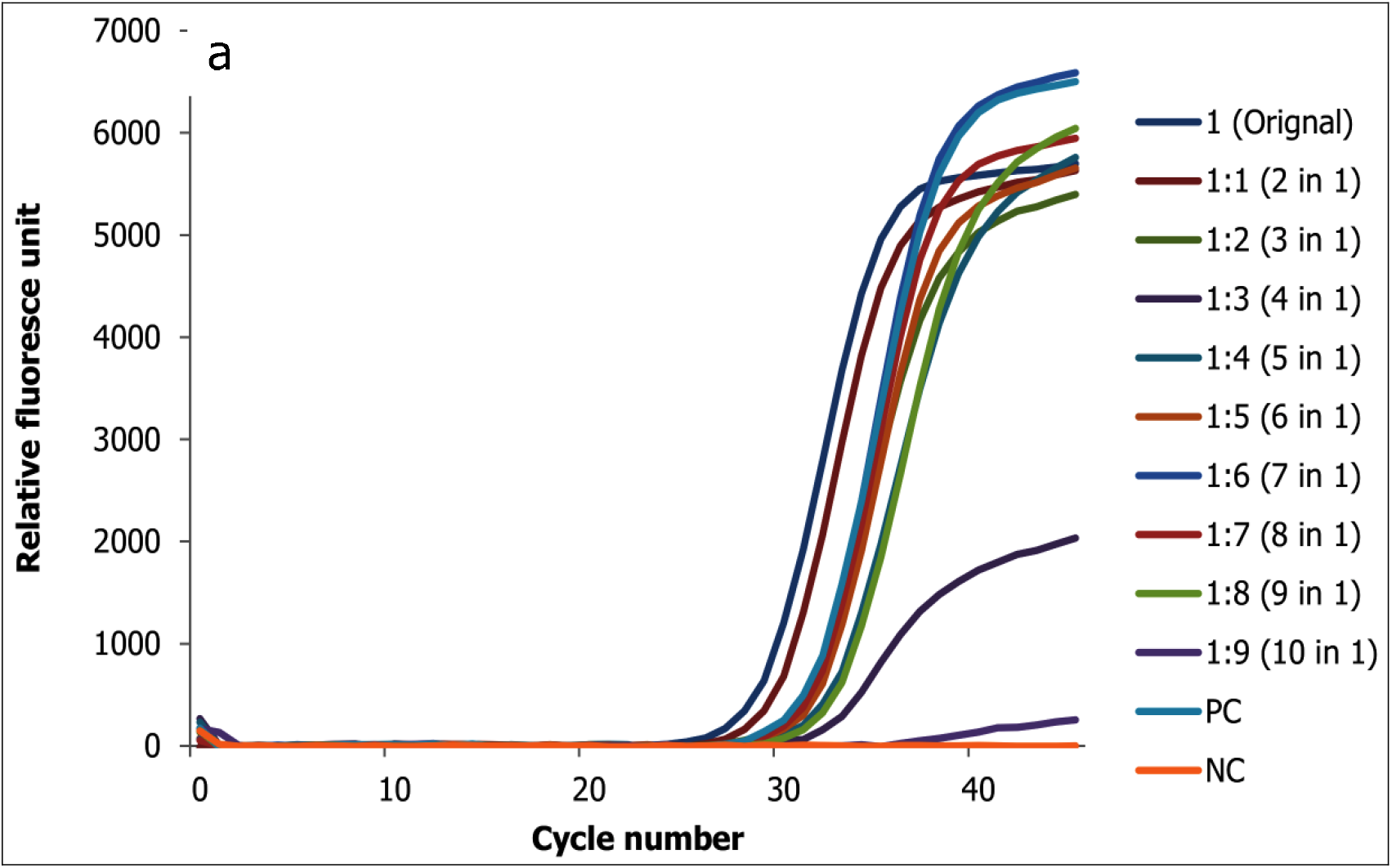
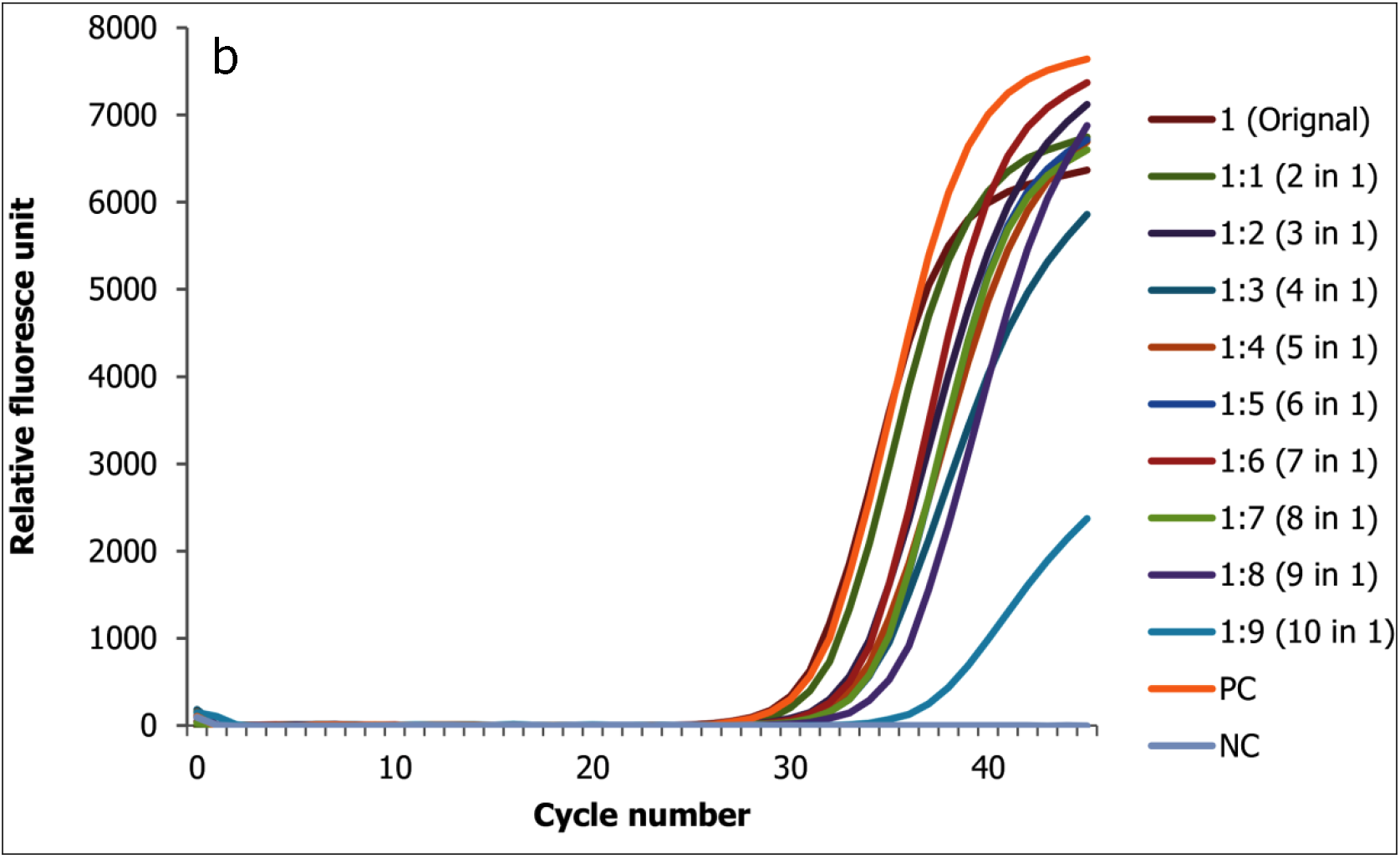
Change in ct value of positive direct biological sample with low ct value (high viral copy) spiked with up to 9 negative samples for the two target genes (1a N and 1b ORF 1ab genes)

Similarly, the SARS CoV-2 positive samples with high ct value (low viral copy number) were within a range of Ct values from 34.37 to 38.82 for N gene and from 37.04 to 39.00 for ORF1ab (data not shown) in 8 in 1pools.

In our RNA pool, we were able to detect SARS-CoV-2 positives samples in pooling up to 10 in 1. Of these, the SARS CoV-2 positive samples with original low ct value (high viral copy number) were within a range of Ct values from 27.74 to 30.93 for N gene and from 29.18 to 32.63 for ORF 1abgene in 10 in 1 pool (Figure 2a and 2b). The results showed that RNA pooled samples were positive within a range of 0 Ct to 3.45 and 0 to 3.19 Ct value difference from the original samples for N and ORF1ab genes, respectively.

**Figure 2**: Change in ct value of RNA positive sample with low ct value (high viral copy) spiked with up to 9 negative samples for the two target genes (2a N gene and 2b ORF1ab gene)

**Figure 2:**
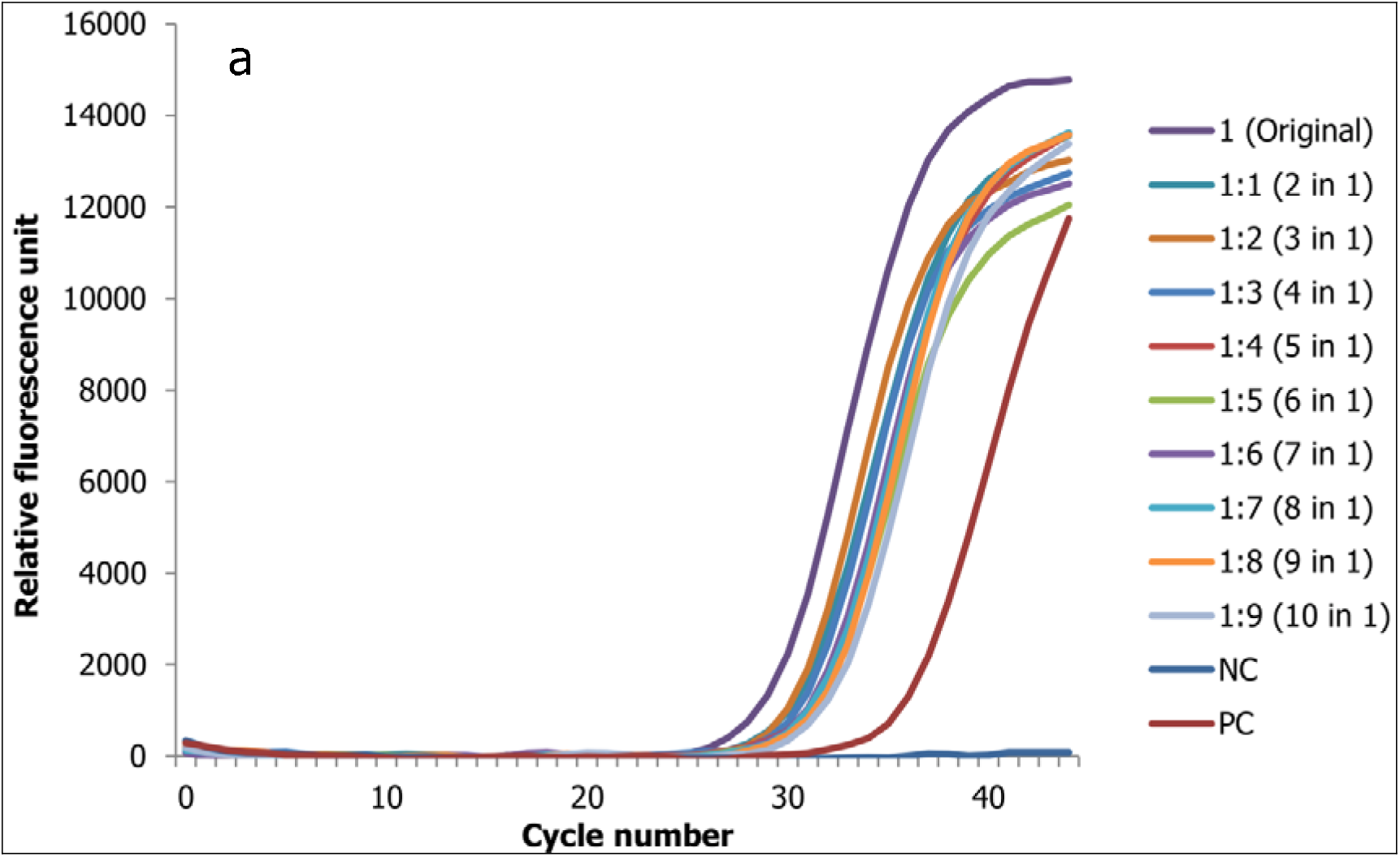
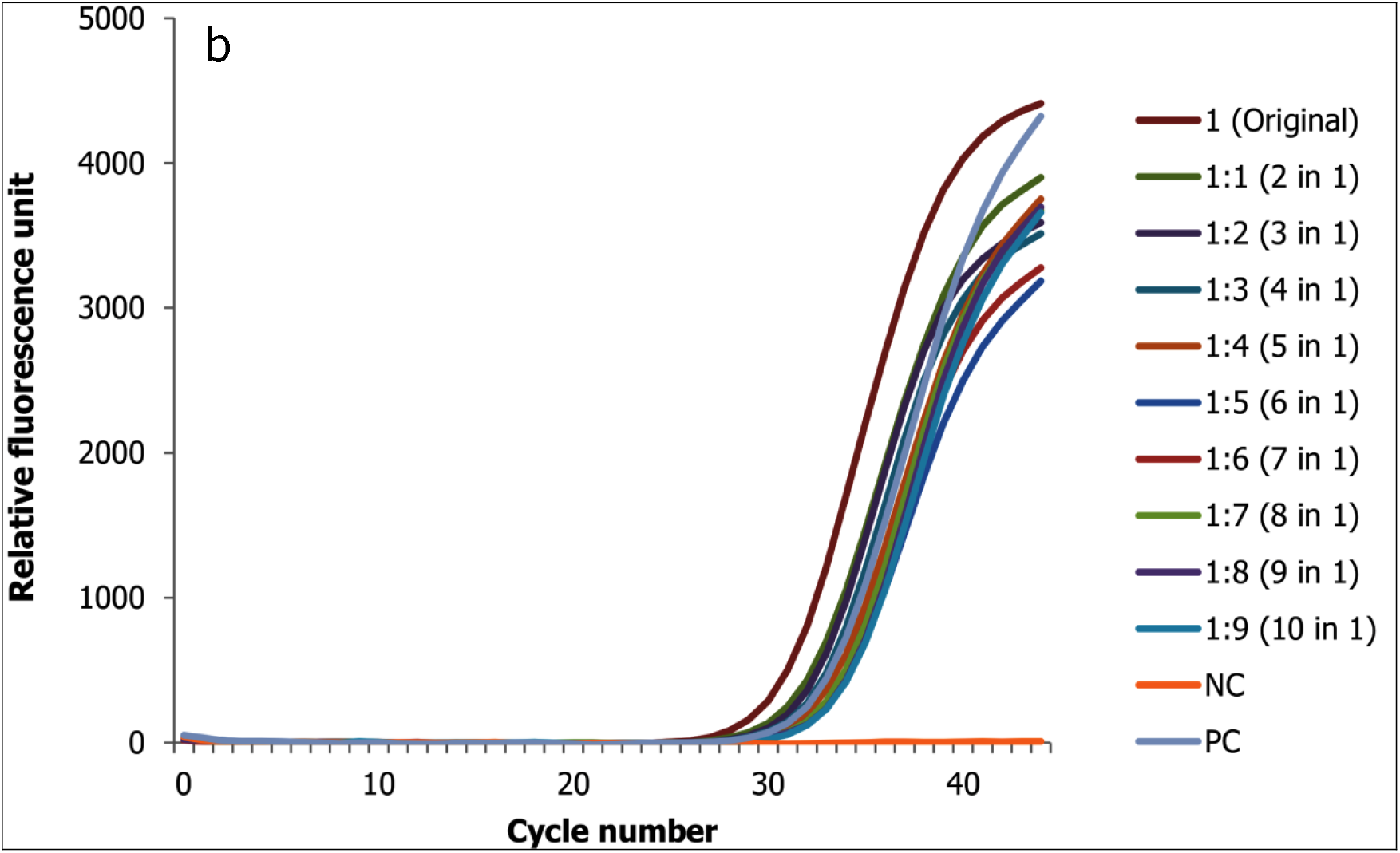
Change in ct value of RNA positive sample with low ct value (high viral copy) spiked with up to 9 negative samples for the two target genes (2a N gene and 2b ORF1ab gene)

Similarly, the SARS CoV-2 positive samples with original high ct value (low viral copy number) were within a range of Ct values from 35.63 to 37.28 for N gene and from 33.91 to 38.39 for ORF1ab in 10 in 1 pool (Figure 3). The results showed that RNA pooled samples were positive within a range of 0 Ct to 3.20 and 0 to 4.48 Ct value difference from the original samples for N and ORF1ab genes, respectively.

**Figure 3:**
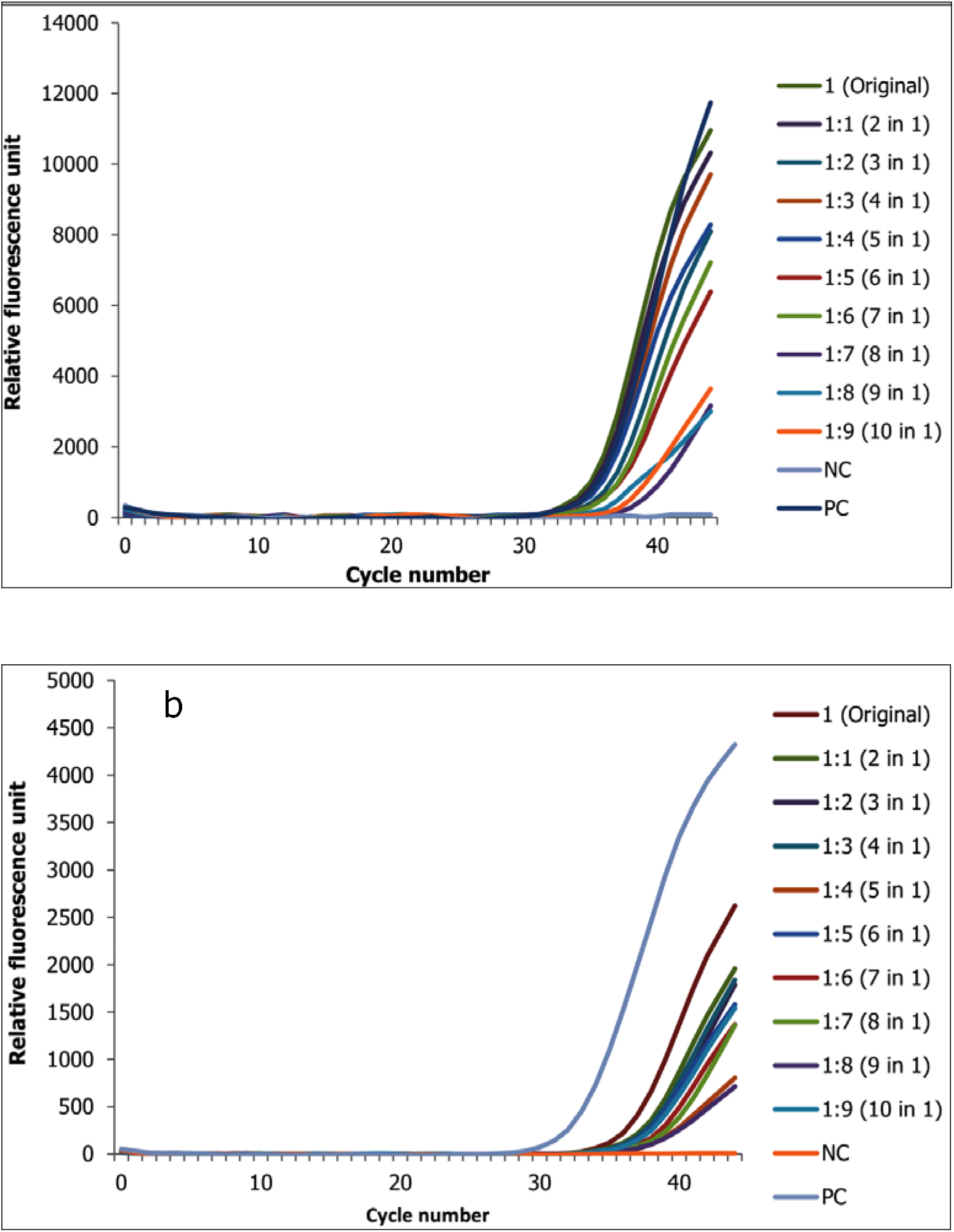
Change in ct value of positive RNA sample high low ct value (low viral copy) spiked with up to 9 negative samples for the two target genes (3a N gene and 3b ORF1ab gene)

As clearly seen in the figures above, as the number of negative pooled samples increases, the amplified RNA reaches the threshold later, as expected from a diluted sample with principle of sample dilution effect. Except for a single replicate (high ct value) which is mixed with 7 negative samples (25 µl of positive and 175 µl of negatives sample from 7 different negative samples), all samples reached the threshold of positivity. For all samples there is a linear correlation between the threshold reached and the doubling of the pool size corresponding with the expectation that an RNA sample that is diluted twice will take one more additional cycle to double. The observed linear correlation indicates that in most cases there is no RNA/DNA (deoxyribonucleic acid) interference with the reverse transcriptase or DNA polymerase enzyme and the amplification process. Of the tested replicates, only replication of high ct value sample did not cross the threshold in pools of 8 (1 positive with 7 negatives). Moreover, with the exception of this specific replicate, the fluorescence of all sample pools increased in a sigmoidal manner. As expected, negative samples in a single or a pool of up to 1:9 samples do maintain no ct values.

## Discussion

Globally, shortage of molecular laboratories for the diagnosis of SARS CoV-2, lack of trained human capital, lack of NA extraction, amplification and detection kits, and lack of accessory and supplementary consumables despite an increasing number of testing demands has become an issue of concern (8, 9) and nationally in a relatively short period of time. To minimize work load, resources and costs, a pooling approach of NA extractions or amplification and detection might be considered. Here, we showed a proof-of-concept for direct clinical sample and RNA/NA pooling for the diagnosis of SARS CoV-2 in Ethiopia using the existing assay.

Results from these pooling methods support that pooled screening strategy can be pursued to increase testing throughput, limit use of reagents, and increase overall testing efficiency [6, 7] at an expected slight loss of sensitivity for direct clinical sample pooling. The same could be attained with no loss of sensitivity for RNA pooling. This study also showed whether pooling was feasible using SARS CoV-2 assay in a both public and clinical setting where the desire to test large numbers of individuals has been impacted by the scarcity of key resources in the country. The predictive algorithm indicated a direct clinical sample pooling ratio of 4 in 1 was expected to retain accuracy of the test irrespective of the ct value (relative RNA copy number) of the sample spiked, and results in a 237% increase in testing efficiency. Furthermore, the predictive algorithm indicated an RNA pooling ratio of 10 in 1 was expected to retain accuracy of the test irrespective of the ct value (relative RNA copy number) of the sample spiked, and results in a 140% increase in testing efficiency.

The practical application of this process was confirmed in the saving of reagents and personnel time that could expand testing. Assuming a consistent positivity rate in the country, this direct biological and RNA pooling strategy would expand testing by 237% and 140%, respectively. However, in a rapidly changing epidemic, testing strategies will need to adapt to real time potential increases in the prevalence rate of a diseases which also requires the use of highly sensitive assays to avoid missing samples with low RNA copy number [7–9]. Furthermore, the impact of different extraction methods on the recovery of RNA/NA and overall assay sensitivity need to be evaluated. And, thus both public and clinical laboratories must perform their own validation pool studies for their own methods of RNA/NA extraction and amplification/detection aligning with the prevalence rates of SARS CoV-2 in real time of the settings. Thus, due to the availability of existing limited SARS CoV-2 diagnosis facility, access to diagnostic tests, kit supplies, the increasing number of individuals to be tested and available trained human capital, this approach facilitates rational use of resources. Furthermore, the approaches could allow for prospective monitoring the effectiveness of contact reduction measures at the population level and early detection of epidemic waves.

However, the limitation of this study is that because of the lack of a plasmid with known concentration, we were not able to quantify the changes occurred in between the dilutions in terms of viral copy number.

## Conclusion

Considering an increasing SARS CoV-2 epidemic and the possibility of unrecognized spread of the diseases within the community we propose a rapid and straightforward screening strategy for SARS CoV-2 using either direct biological sample polling 4 in 1 or RNA pooling up to 8 in 1. This approach proved its concept and principles, and may facilitate detection of early community transmission of SARS CoV-2 to enable the timely implementation of appropriate infection control measures to reduce spread. The method can also be used for routine monitoring of essential work groups.

## Data Availability

All data generated or analysed during this study are included in this article

## List of abbreviations

ct: Cycle threshold
COVID-19: Coronavirus disease 2019
DNA: Deoxyribonucleic acid
E: Envelop
LOD: Limit of detection
MM: Master Mix
µl: Micro liter
NA: Nucleic acid
ORF1ab: Open reading frame
RT-PCR: Reverse transcriptase polymerase chain reaction
RNA: Ribonucleic acid
SARS CoV-2: Severe acute respiratory syndrome coronavirus 2

## Declarations

### Ethics

Being an Internal Experiment at methods validation level and given the decoded nature of testing, individual patient consent was not required and conducted based on institutional system. And, the Armauer Hansen Research Institute/ALERT Ethics Review Committee has also waived it.

### Consent to publication

Not applicable

### Availability of data and materials

All data generated or analysed during this study are included in this article

### Competing interests

The authors declare that they have no competing interests

### Funding

None

### Authors’ contributions

AM: Conceptualization, Formal analysis, Methodology, Supervision, Writing – review & editing

DAH: Investigation, Formal analysis, Methodology, Writing – review & editing

FA: Investigation, Formal analysis, Methodology, Writing – review & editing

DAT: Investigation, Formal analysis, Methodology, Writing – review & editing

SW: Investigation, Formal analysis, Methodology, Writing – review & editing

GA: Investigation, Formal analysis, Methodology, Writing – review & editing

TS: Investigation, Formal analysis, Methodology, Writing – review & editing

MH: Investigation, Formal analysis, Methodology, Writing – review & editing

AA: Investigation, Formal analysis, Methodology, Writing – review & editing

AGB: Investigation, Formal analysis, Methodology, Writing – review & editing

GTB: Investigation, Formal analysis, Methodology, Writing – review & editing

## Acknowledgements

Not applicable

## Notes

### Competing Interest Statement

The authors have declared no competing interest.

